# Analyses of health surveys indicates regions of priority to eliminate open defecation in Africa and implication for antimicrobial resistance burden

**DOI:** 10.1101/2023.07.21.23293022

**Authors:** Omololu Ebenezer Fagunwa, Thobile Mthiyane, Ayokunle Fagunwa, Kassim Idowu Olayemi, Alaoma Alozie, Helen Onyeaka, Adenike Akinsemolu, Adegbola Ojo

## Abstract

Sanitation, which offer safe and effective methods for waste disposal is important for development. However, in Africa and other developing regions, the prevalent practice of open defecation (OD) impedes attaining the Sustainable Development Goals (SDGs). This research delves into the analysis of OD in Africa and proposes a three-tier priority system, comprising critical, high, and medium areas, through which developmental endeavours can be targeted. To achieve this, the study utilizes data from Demographic and Health Surveys (DHS) and World Bank. The rates of OD at country and sub-country/region levels were calculated to define the priority system and regression analyses were used to determine predictors of OD practice. The findings are that Nigeria, Ethiopia, Niger, the Democratic Republic of Congo, Burkina Faso, and Chad have a high number of people struggling with open defecation. In addition, disparities in access to proper sanitation facilities were identified among impoverished individuals and those residing in rural areas. After adjusting for education and residence, the poorest are 43 times (95% confidence interval *42.443 – 45.290*) more likely to practice open defecation in comparison to the wealthiest. Consequently, wealth index is a pivotal factor in eradicating open toileting. To address this pressing issue in Africa, it is imperative to prioritize evidence-based targeted interventions that concentrate on regions and communities urgently needing improved sanitation infrastructure and programmes. Special attention should be paid to West Africa since many of its communities are in the critical category. Poverty and inequality must be addressed and investments in sanitation infrastructure, behavioural change promotion, and support for multistakeholder collaborations should be encouraged. To evaluate OD intervention and monitor health impact, variables such as antimicrobial resistance (AMR) should be included in important health surveys (e.g., DHS). This study is the largest meta-data analyses of OD in Africa detailing drivers and communities that should be prioritised on sanitation interventions.

## INTRODUCTION

The United Nations (UN) recognizes access to clean water, sanitation, and health (WASH) as a human right. Despite progress in improving access to these essential services, billions of people still lack them. According to the UN (2015) and United Nations Development Programme (UNDP) (2021), over 4.5 billion people or over half of the global population lack safe sanitation. Additionally, 946 million people continue to practice open defecation (OD), which is the act of disposing of human waste in open areas rather than using a toilet.

Economic and cultural belief systems drive OD practices (Kar and Milward, 2011; Ntaro *et al.,* 2022; Gauri *et al.,* 2023). Public health officials must understand the complex interactions between socio-economic, behavioural, and environmental factors to combat this practice and develop context-specific strategies (Giribabu *et al.,* 2019). One successful approach is Community-led Total Sanitation (CLTS), which has contributed to significant progress in the Millennium Development Goals across Asia and Sub-Saharan Africa. In CLTS, communities appraise and analyse OD, then mobilise people to identify solutions and monitor progress (Kar and Milward, 2011; Kouassi *et al.,* 2023). OD is a threat to human dignity, poses environmental and health risks (Giribabu *et al.,* 2019) and is linked to a significant number of deaths from cholera and typhoid in developing countries (Mara, 2017; Akilimali *et al.,* 2023).

OD is a degrading, polluting, and dangerous practice that disproportionately affects developing countries, particularly in Asia and Sub-Saharan Africa. 90% of people who engage in OD reside in rural areas of these regions (WHO/UNICEF, 2017; Saleem *et al.,* 2019; UN DESA, 2021). The United Nations department of economic and social affairs (2021) noted that OD clearly indicates extreme poverty in affected countries. Studies have shown that improved sanitation can reduce the transmission of enteric pathogens and intestinal parasites, decreasing morbidity and mortality, especially in children (Mara, 2017). Therefore, the Sustainable Development Goals (SDGs) offer a holistic solution to the social, health, and environmental challenges posed by OD by maximizing synergies and collaborating to address water and sanitation issues rather than relying on limited individual interventions.

### The Sustainability Goals on Water and Sanitation

There has been deliberate effort and interventions since the beginning of this Millennium to end OD to promote good health and a sustainable environment and, therefore, the basis of the target 7C of the Millennium Development Goals (MDGs) relating to sustainable access to safe drinking water and basic sanitation (WHO, 2018). SDG 6 aims to expand access to basic water and sanitation services and close the gaps in service quality. The SDG 6 has six outcome oriented targets but with a total (including sub-targets of 8) of 11 indicators to represent the metrics for tracking the achievement of the targets, of which target 6.2 appears to be a rebirth of the MDG target 7C. Notably, a total of 2.6 billion people gained access to safe water in the MDG regime, and 2.1 billion obtained access to safe sanitation (United Nations, 2015). Despite the success achieved, criticism emerged regarding the ambiguities of the classification “improved and unimproved” as used in the report presented. However, this was subsequently reviewed and addressed by the WHO/UNICEF Joint Monitoring Programme (JMP) 2017 report. This paper does not intend to engage in the dialectics or semantics to argue the operationalisation of these constructs but to make a case for retrospective understanding, analyses of current status and future thinking to be able to develop robust and pragmatic context-relevant measures that can sever these societies from the vulnerable practice of OD. At the beginning of the millennium, a non-profit organisation, World Toilet Organisation, was formed to draw global attention to the sanitation crisis and established November 19 as world toilet day (WTO, 2021). In less than a decade, the international year of sanitation was commemorated to encourage countries to commit to increasing safe toilet facilities and ending open defecation (UN Water, 2008). The fifteen-year implementation phase of MDGs ended without any country in sub-Saharan Africa achieving target 7C; however, the SGD 6 holds a promise that changes the story.

### Clean Water and Sanitation

The sixth goal of the SDGs is focused on providing access to clean water and sanitation. While an estimated 5.2 billion people had access to safely managed drinking water in 2015, 844 million still lacked basic access. Similarly, while 2.9 billion people had access to safe sanitation in 2015, 2.3 billion people still lacked basic sanitation facilities (UNDP, 2021). These challenges are further compounded by the fact that 892 million people still practice open defecation (CDC, 2022). Target 6.2 is that by 2030, the world should achieve access to adequate and equitable sanitation and hygiene for all and end open defecation, paying particular attention to the needs of women, girls, and those in vulnerable situations (UNDP, 2021). In addition, the issue of water security, a significant factor in sanitation and a critical resource in the effort to end OD is a priority area in Agenda 2063, the seventh goal of the African Union strategic framework aligns with SDG6 (AU, 2015; 2021). This highlights global and regional efforts to halt many of the problems developing countries face and free them from the constraints of poverty and its manifestations. The essentiality of water implies that it touches every aspect of development and is a critical nexus for nearly every SDG. Recently, the US International Development Association (IDA), through the World Bank, approved a $700m credit to Nigeria to pursue the Sustainable Urban and Rural Water Supply, Sanitation, and Hygiene Programme (SURWASH) that will provide 6 million people with basic drinking water services and 1.4m people access to improved sanitation services with the potential to support 500 communities to achieve open defecation free status (Nasir, 2021).

#### Why toilet and burden of open defecation

Toilet is a crucial development amenity since it positively correlates with achieving other SDGs. For instance, a clean and safe toilet enhances educational attainment (SDG4) and reduces gender inequalities (SDG5) (Unterhalter *et al.,* 2014; Daniel *et al.,* 2023; Das *et al.,* 2023). Similarly, investment in sanitation has been reported to generate a quantifiable, positive return on investment with savings on medical costs and increased productivity. The assessment paper established a relationship between sanitation and economic growth (SDG8). It posed that every $1 invested in basic sanitation yields a return of $3 (Hutton *et al.,* 2015; UN Water 2023). The more obvious importance of a clean and safe toilet relates to attaining good health and wellbeing. Inadequate drinking water, sanitation, and hygiene are important risk factors, especially in low-and middle-income countries (WHO, 2022). A systematic review of sanitation studies published between 1970 to 2013 indicates that about 280,000 diarrhoea deaths occurred due to inadequate sanitation with figures increasing yearly (Prüss-Ustün *et al.,* 2014; 2019). OD encourages the transmission of pathogens (such as bacteria and viruses) and parasites (such as nematodes and helminths) which cause damage to human health (Carr, 2001; Dandabathula *et al.,* 2019; Prüss-Ustün *et al.,* 2019). OD also affect production and food safety. Salmonella, usually present in human faeces, negatively affects maize and bean seeds (Singh *et al.,* 2007). In addition, it can also lead to direct and indirect food safety issues. For example, flies and other insects are attracted to faeces. They can carry bacteria from faecal matter to food, leading to contamination. Animals may also be attracted to faeces, which can lead to the contamination of food sources through direct contact or the spread of faeces-borne diseases (Guthami *et al.,*2017). Open defecation contributes to the contamination of soil and water, and the transmission of diseases (Joab *et al.,*2017; Miranda *et al.,*2018; Brooks *et al.,* 2023). A global assessment of faecal exposure found that contamination of water sources by pathogens through faeces is highest in Africa and add more pressure on the efforts to close the gap on health disparity (Bain *et al.,* 2014). More worrisome is that there are well established evidence showing that antimicrobial resistance (AMR) pathogens developed and spread through open defecation and sub-Saharan Africa has the highest AMR burden (Wellcome Trust *et al.* 2018; Hendriksen *et al.,* 2019; Murray *et al.,* 2022).

The high population growth rate in Africa, especially in Sub-Saharan, affects attaining an open defecation-free continent (Abebe and Tucho, 2020). A study reported that among 34 Sub Saharan African countries, only Angola, Ethiopia and Sao Tome and Principe had a ≥10% reduction in OD between 2005-2010 (Galan *et al.,* 2013). The provision of functioning latrine toilets has been recognised as a cost-effective way to end open defecation. This has been an implementation focus during the MDGs regime (Tyndale-Biscoe, Bond and Kidd, 2013; Kipkoech *et al.,* 2023). However, the provision of toilets does not always equate to the end of open defecation (Singh and WSP, 2007; Sinha, 2019). Even when the provision of toilets is assumed to be zero OD, a continental study reported that 13% of the beneficiaries reverse into practising OD after about two years and the same elsewhere in Asia (Tyndale-Biscoe, Bond and Kidd, 2013; Augsburg *et al.,* 2022). Financial support, lack of follow-up support, inconvenience, discomfort, sharing with others, maintenance, and repair were identified as reasons for the return to OD or slippage.

### Determinants of open defecation

The threat of OD practice called for an understanding its determinants. One of the earliest efforts in WASH is the development of a theoretical framework called FOAM – Focus on Opportunity, Ability and Motivation (Coombes and Devine, 2010; Khare and Suresh, 2021). The framework was designed to support WASH implementers in the development, evaluation, and monitoring of behavioural change. Targeting population is recognised within the ’focus’ stand of the framework and well adapted to in understanding the determinant of open defecation. The variables that are linked to OD differs depending on population of studies (Abubakar, 2018). Notwithstanding, the most identified determinants of OD include education, occupation, area of residence, wealth, farming distance from home, weather and seasons, use of substances and alcohol, household size, gut health status and enteric diseases, government policies and investment, geopolitical region, ownership of toileting facilities (Abubakar, 2018; Osumanu *et al.,* 2019; UN Water, 2020; Belay *et al.,* 2022; Ali and Khan, 2023).

#### A Pan African Study on Open Defecation

The MDGs report states that the proportion of the global rural population practising open defecation has fallen from 38 % to 25% between 1990 – 2015 (UN, 2015). However, using a global aggregate, the MDG report masked the contextual realities of African countries. Many African countries are far from the pathway to ending OD. This appears to play down the seriousness of the problem and tackle it head-on within the next SDGs phase. Reports show that most countries lagging in meeting the access to adequate sanitation targets are located in Sub-Saharan Africa (Waage *et al.,* 2010; UNICEF and WHO, 2020). Although there are efforts through policy-driven agendas in continental Africa and at regional levels for example the African Union Agenda 2063 and ECOWAS vision 2050 to end open defecation by 2030 under the current SDGs, there is a lack of pragmatic efforts with regards to funding and implementation (NEPAD, 2022; ECOWAS, 2022). Yet, approximately 60 million Nigerians lacked access to basic drinking water services, 80 million lacked improved sanitation, and 167 million lacked a basic handwashing facility. In rural regions, 39% of families lack access to at least basic water supply services. At the same time, only half of this number have access to improved sanitation and about a third practise open defecation. This percentage has remained relatively unchanged since 1990 (Nasir, 2021). In Kenya, its bureau of statistics demonstrates that only 39% of Kenyans use unimproved sanitation facilities (Busienei, Ogendi and Mokua, 2019). There are over 25 African countries with more than 15% of their population practising OD (WHO, 2015; WHO/UNICEF JMP, 2021), and this is as high as 73% in Niger, 74% in South Sudan and 77% in Eritrea. Studies that synthesize multinational large datasets on OD and capture diverse range of contexts is still lacking. This study aims to evaluate prevalence and sociodemographic drivers of open toileting in Africa, and highlights communities in urgent need based on three-tier priority system. These will allow for addressing the contextual realities of African countries and the need for pragmatic and efforts to tackle the persisting OD issue.

## METHODS

### Study Population

The study population for this research was primarily drawn from Demographic and Health Survey (DHS) data for African countries. The data collection for DHS was approved by the ethics committee of each host country, consisting of the statistics office/bureau and health ministry. Technical support for the survey was given by ICF International Inc. The DHS program surveys are nationally representative. DHS program office in Rockville, Maryland, approved using de-identified datasets from African countries for analysis of water, sanitation, and hygiene.

Data used in this study were available acquired immunodeficiency syndrome (AIDS) indicator survey (AIS), malaria indicator survey (MIS) and demographic and health surveys (DHS). Two hundred and twenty-two health surveys across Africa were checked for sanitation variables. The total sample size is 8,659,881. All these surveys are nationally representative population based surveys. However, AIS focus on human immune-deficiency virus /AIDS and related issues, while the MIS focus on malaria and associated issues, with a sample of about 3,000 households each. The DHS have a larger sample size, usually 5,000 to 30,000 households. Toileting questions used throughout this study are under household member recode. The latest DHS phase available for a country is chosen as the most recent data available. The same is used to depict the current situation. For instance, Angola has datasets AOPR51FL, AOPR62FL, and AOPR71FL. Still, dataset AOPR71FL is the most recent dataset from this country since it was conducted in phase 7. When MIS, AIS, or DHS datasets are available in the same phase in a country (e.g., Ghana), the standard DHS dataset is used instead since it is more extensive. Where AIS and MIS only are available in the most recent phase, the dataset with a later collection date is chosen (e.g., Mozambique). The total sample size is 8,659,881, the datasets used were sourced from DHS (DHS, 2022). Eastern and Western African surveys have the highest number (Table 1).

**Table 1:** The forty African countries showing sample size and regions.

| **Overview of datasets used** |  |  |  |  |  |
| --- | --- | --- | --- | --- | --- |
| Country | Sample size | No of Surveys | Country | Sample size | No of Surveys |
| **Central** |  |  | **Eastern** |  |  |
| Angola | 129893 | 3 | Burundi | 144073 | 3 |
| Cameroon | 171652 | 4 | Comoros | 38631 | 2 |
| Central Africa Republic | 27992 | 1 | Ethiopia | 289033 | 4 |
| Chad | 165899 | 3 | Kenya | 331483 | 6 |
| Republic of Congo | 112562 | 3 | Madagascar | 319088 | 7 |
| Democratic Republic of Congo | 144071 | 2 | Malawi | 373553 | 8 |
| Gabon | 73973 | 2 | Mozambique | 258930 | 6 |
| Sao Tome and Principe | 13430 | 1 | Rwanda | 309542 | 7 |
| **Western** |  |  | Tanzania | 450922 | 10 |
| Benin | 311612 | 5 | Uganda | 358252 | 8 |
| Burkina Faso | 284969 | 6 | Zambia | 230934 | 5 |
| Cote d'Ivoire | 127768 | 4 | Zimbabwe | 186845 | 5 |
| The Gambia | 52647 | 1 | **Northern** |  |  |
| Ghana | 184836 | 6 | Egypt | 619789 | 7 |
| Guinea | 166434 | 4 | Morocco | 104007 | 2 |
| Liberia | 147027 | 5 | **Southern** |  |  |
| Mali | 342182 | 6 | Lesotho | 125169 | 3 |
| Niger | 182768 | 4 | Namibia | 142373 | 4 |
| Nigeria | 627623 | 7 | South Africa | 91428 | 2 |
| Senegal | 720890 | 13 | Swaziland<br>(Eswatini) | 26974 | 2 |
| Sierra Leone | 157170 | 3 |  |  |  |
| Togo | 113457 | 3 |  |  |  |

### Assumptions for 2030 OD projections

To understand the current trend of the top ten African countries practising open defecation based on population, statistics from the World Bank population estimates for the year 2020 were used. Similarly, a 2030 projection on how challenging OD may be in these ten countries made use of the World Bank population projection for the year 2030 (World Bank, 2023).

**Country and regional grouping**: 39 African countries in the study are grouped into five sub regional categories. These categories follow the United Nations M49 standard classification of countries worldwide (UNSD, 1999). The type of data included for each sub-region and country is presented in the supplementary material.

### Survey questionnaires, variables and covariates

The DHS program uses standard model questionnaires and a written description of why specific questions have been included. The current and previous versions of questionnaires used for the DHS, MIS, and AIS are open source at DHS Program methodology section. The dependent variable used in this study is ’*type of toilet facility’* HV205. No facility/outdoor/bush =31 was used for open defecation. Other values indicate using a type of toilet, covering various flush toilets and latrine/pit toilet options. At the same time, other variables are *’type of place of residence’* HV025, *’region/province’* HV024, and *’wealth index’* HV207. Known underlying factors and determinants that may contribute to open defecation were explored. The determinants probed are poverty, sex, age group, place of residence, and education. These determinants were included as covariates in the analysis.

### Regression analysis

Multinominal regression analysis was performed for regions with critical priority regions. The dependent variable *’type of toilet’* HV205 was recoded using three categories: Flush toilet (values 10-19), Latrine toilet (values 20-29) and No toilet/Open defecation (values 30-31). The independent variables include *’wealth index combined’* HV270 as a factor. Covariates are *’highest education level attained’* HV106 and ’*type of place of residence*’ HV025.

The equation used for multinomial regression analysis is:

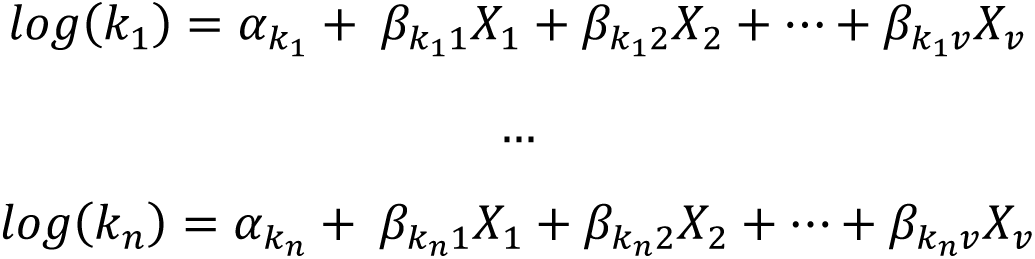

Where *log*(*k_n_*) is the natural logarithm of class *k_n_*versus the reference class *k*_0_, *X* is a set of explanatory variables (*X*_1_, *X*_2_ … . *X_v_*), *k_n_* is the intercept term for class *k_n_* versus the reference class, and *β* is the slopes for the classes.

The equation for binomial regression analysis is:

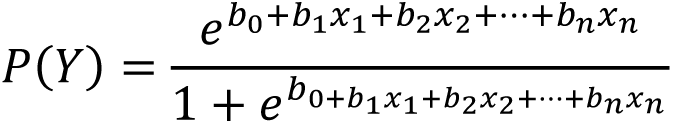

Where *P* the probability of *Y* occurring, *e* is the natural logarithm base, *b*_0_is the intercept at the y-axis, *b*_1_ is the line gradient, *b*_n_is the regression coefficient of *X*_n_, *X*_1_ is the predictor variable. The minimum variable is two. (Chatterjee and Simonoff, 2013).

### Priority categorisation

To see where there is a need for intense sanitation programmes and efforts, regions were classified into three categories:

Critical priority (≥80%), for geographical locations where the open defecation rate is greater than 80%.

High priority (60-≤80%), for geographical locations where the open defecation rate is between 60% to 80%.

Medium priority (40-≤60%), for geographical locations where the open defecation rate is between 40% and less than 60%.

### Ethics statements

For this study, no ethical approval was needed from individuals or participants because it was a secondary data analysis of de-identified data, originally collected by the DHS Program and generously made available. DHS Program approved the use of the datasets.

## RESULTS

The estimated rate of open defecation by population revealed that Nigeria (54 million), Ethiopia (43 million) and Niger (15 million) top the table of African countries with the highest number of people practising OD (Table 2). Others include DR Congo, Burkina Faso, Chad, Angola, Madagascar, Kenya, and Cote d’Ivoire. These ten countries could account for about 247 million Africans defecating in the open by 2030 if critical and emergency actions are not taken. The datasets used for analysis were from DHS Program and the World Bank (DHS, 2022; World Bank, 2023).

**Table 2:**
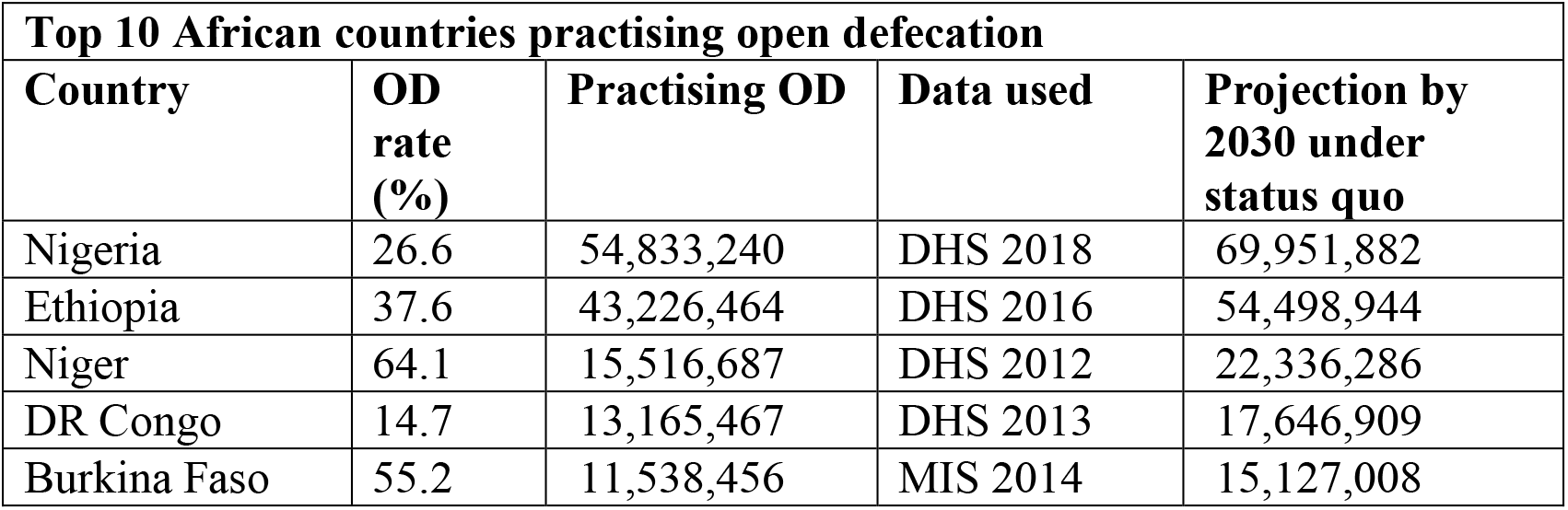
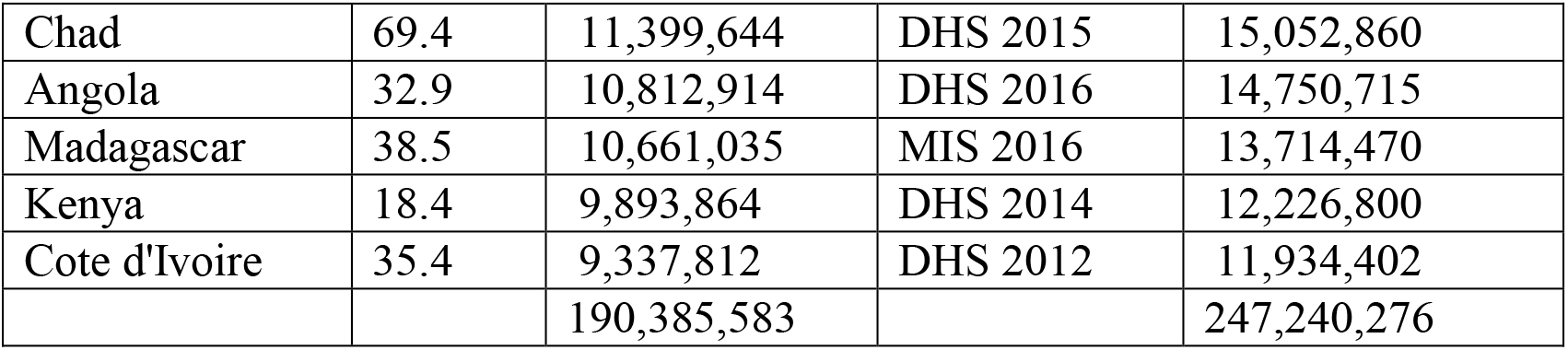
Top ten African countries practising open defecation based on population.

| **Top 10 African countries practising open defecation** |  |  |  |  |
| --- | --- | --- | --- | --- |
| **Country** | **OD rate (%)** | **Practising OD** | **Data used** | **Projection by 2030 under status quo** |
| Nigeria | 26.6 | 54,833,240 | DHS 2018 | 69,951,882 |
| Ethiopia | 37.6 | 43,226,464 | DHS 2016 | 54,498,944 |
| Niger | 64.1 | 15,516,687 | DHS 2012 | 22,336,286 |
| DR Congo | 14.7 | 13,165,467 | DHS 2013 | 17,646,909 |
| Burkina Faso | 55.2 | 11,538,456 | MIS 2014 | 15,127,008 |
| Chad | 69.4 | 11,399,644 | DHS 2015 | 15,052,860 |
| Angola | 32.9 | 10,812,914 | DHS 2016 | 14,750,715 |
| Madagascar | 38.5 | 10,661,035 | MIS 2016 | 13,714,470 |
| Kenya | 18.4 | 9,893,864 | DHS 2014 | 12,226,800 |
| Cote d'Ivoire | 35.4 | 9,337,812 | DHS 2012 | 11,934,402 |
|  |  | 190,385,583 |  | 247,240,276 |

Analyses of the past DHS datasets indicate that Egypt, Comoros, and Rwanda are historically on the right path to ending open defecation (Table 3). Just only 0.1% of the population of Egypt practices open defecation. Within fourteen years, the country went from a 3.5% rate to 0.1%. Egypt is one of African most feasible countries to end open defecation by 2030. Historically, nearly Open defecation-free countries include Egypt, Comoros, Rwanda, Burundi, Gabon, Gambia, and South Africa. The datasets were from the DHS Program (DHS, 2022).

**Table 3:** Countries that historically have <5% open defecation rate (green flag). Rows in the shade indicate the most current open defecation rate.

| **Low open defecation rates in history** |  |  |  |  |
| --- | --- | --- | --- | --- |
| **Country** | **Year of Study** | **Data** | **Code** | **OD rate (%)** |
| Egypt | 2000 | DHS | EGPR42FL | 3.5 |
|  | 2003 | DHS | EGPR4AFL | 2.0 |
|  | 2005 | DHS | EGPR51FL | 1.6 |
|  | 2008 | DHS | EGPR5AFL | 0.5 |
|  | 2014 | DHS | EGPR61FL | 0.1 |
| Comoros | 1996 | DHS | KMPR32FL | 0.3 |
|  | 2012 | DHS | KMPR61FL | 0.7 |
| Rwanda | 2000 | DHS | RWPR41FL | 2.9 |
|  | 2005 | DHS | RWPR53FL | 3.3 |
|  | 2008 | DHS | RWPR5AFL | 2.0 |
|  | 2010 | DHS | RWPR61FL | 1.0 |
|  | 2013 | MIS | RWPR6QFL | 1.7 |
|  | 2015 | DHS | RWPR70FL | 2.9 |
|  | 2017 | MIS | RWPR7AFL | 1.4 |
| Cameroon | 1991 | DHS | CMPR22FL | 2.1 |
| Burundi | 2010 | DHS | BUPR61FL | 2.6 |
|  | 2012 | MIS | BUPR6HFL | 2.2 |
|  | 2017 | DHS | BUPR70FL | 1.9 |
| South Africa | 2016 | DHS | ZAPR71FL | 2.6 |
| Gabon | 2000 | DHS | GAPR41FL | 2.7 |
|  | 2012 | DHS | GAPR60FL | 2.5 |
|  | 2012 | DHS | GAPR60FL | 2.5 |
| Gambia | 2013 | DHS | GMPR60FL | 2.5 |
| Malawi | 2014 | MIS | MWPR71FL | 4.2 |
|  | 2015 | DHS | MWPR7AFL | 5.2 |

Countries that historically have OD rate >50% are Benin, Ethiopia, Niger, Burkina Faso, Togo, Chad, Namibia, Sao Tome and Principe, Madagascar, Liberia, and Mozambique (Table 4). These countries have always been highly burdened with sanitation challenges. More than half of the population in these countries still has no toilet facilities except for Mozambique (21.7%), Madagascar (38.5%), Liberia (40.5%), and Burkina Faso (42.7%). The datasets were from the DHS Program (DHS, 2022).

**Table 4:**
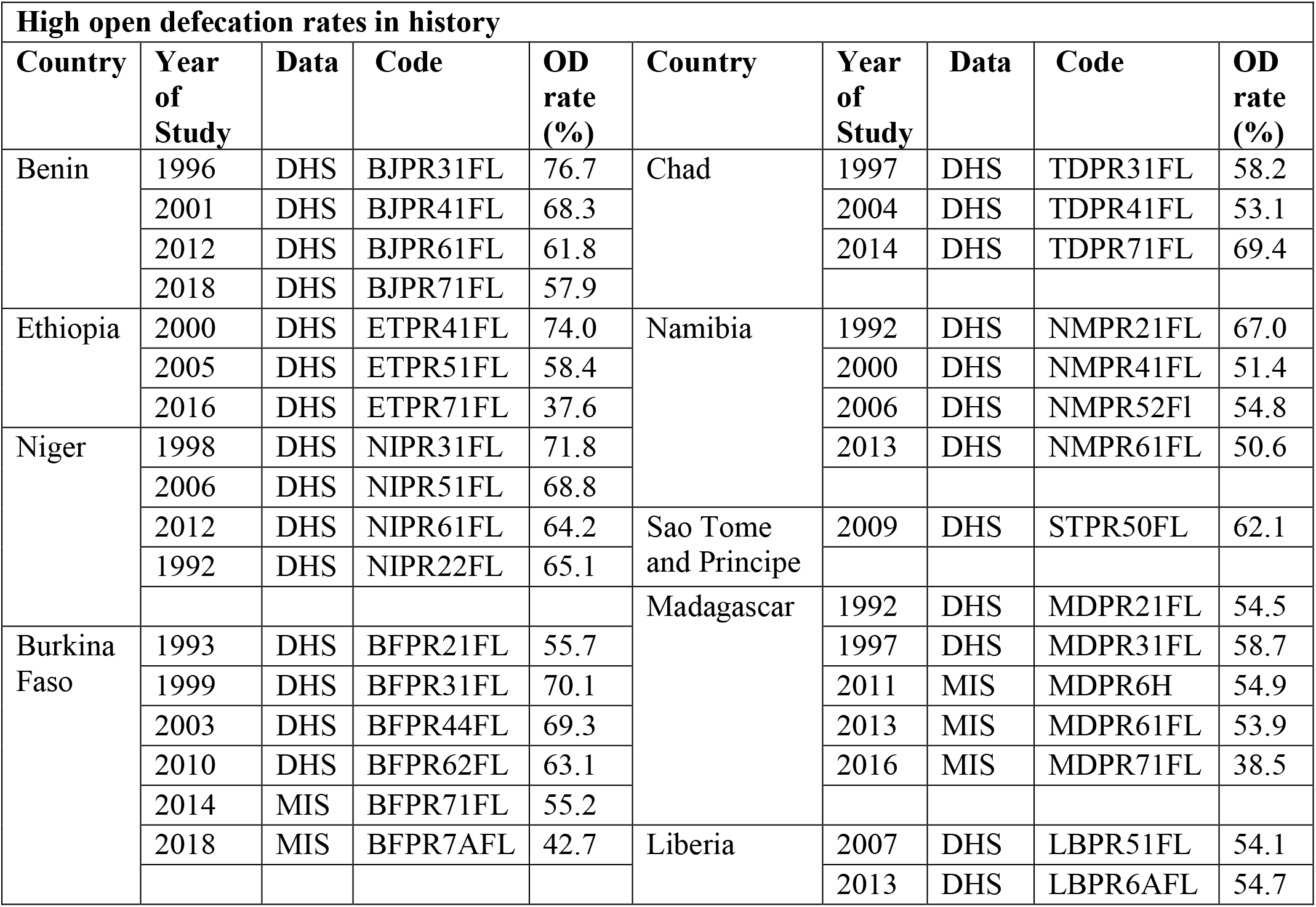
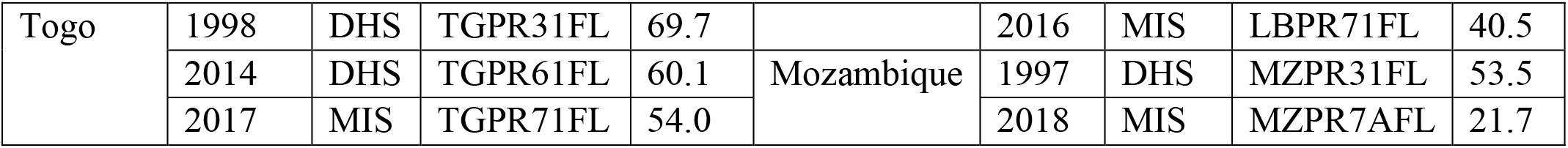
Countries that historically have >50% open defecation rate (red flag).

The sub-national/regional/provincial level burden indicates that it is still high in some African urban areas (Figure 1). Benin, Chad, Namibia, and Sao Tome and Principe particularly have the challenge of sanitation in their urban centres. Provinces with a critical urban OD rates are *Atacora* (81.9%), and South region (80.9%) in Benin and Sao Tome and Principe, respectively. The high-priority regions are *Ennedi* and *Lac* (Chad), *Karamoja* (Uganda), *Est* (Burkina Faso), and *Ohangwena* (Namibia), with OD rate of 73.1%, 65.0%, 68.1%, 65.0%, and 63.9% respectively. The thirty-five regions requiring urgent action comprise two critical priority regions, five high-priority regions and 28 medium-priority regions. The datasets were from the DHS Program (DHS, 2022).

**Figure 1:**
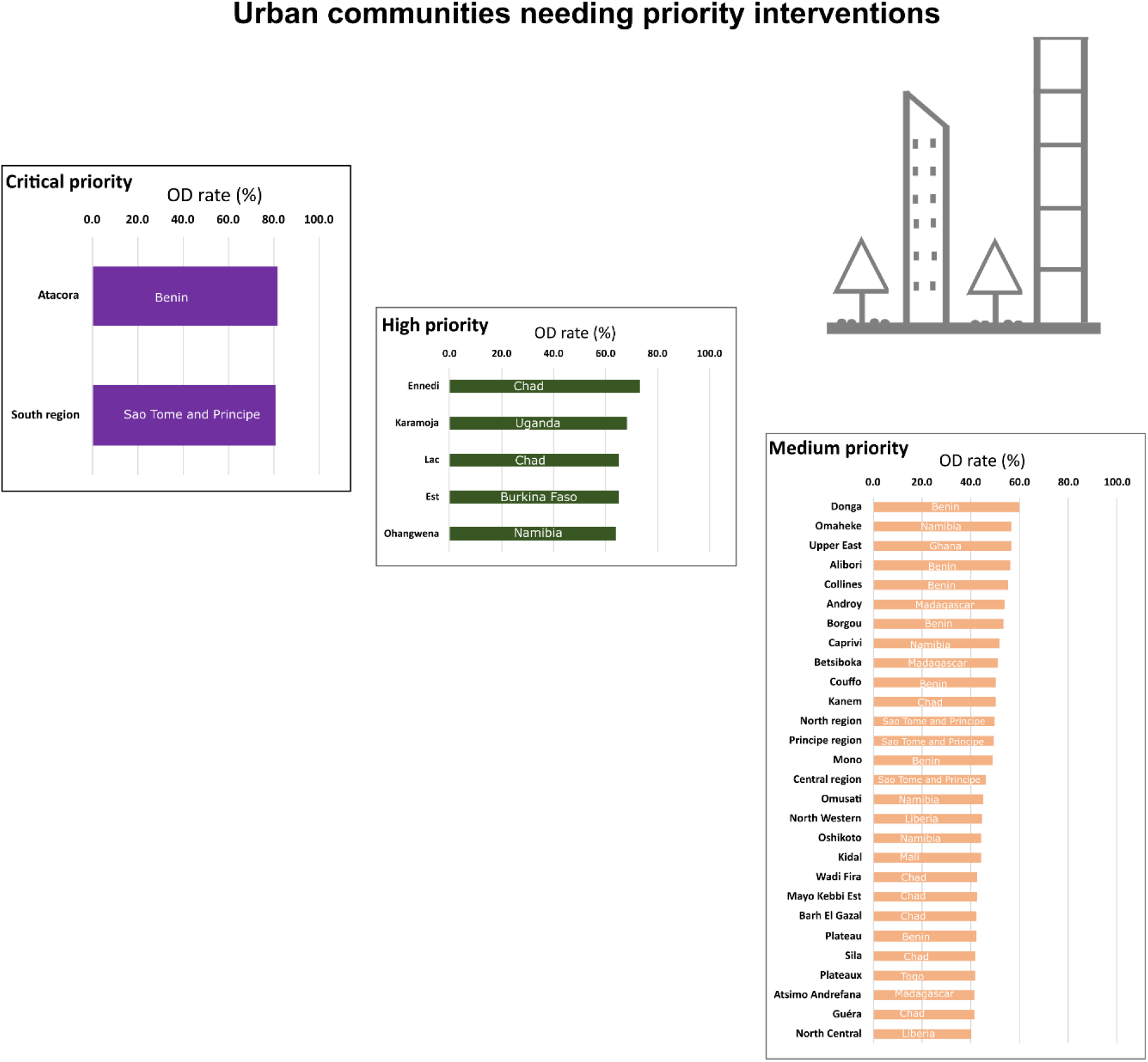
Urban communities in Africa needing priority interventions against open defecation.

Figure 2 indicates that OD is mainly at a critical or high-priority state in rural areas of Africa. The burden at the sub-national level means that Lac (98.7%), Kanem (97.3%), Ouaddaï (96.9%), Batha (92.5%), Barh El Gazai (91.8%), Logone Oriental (80.2%), Borkou/Tibesti (89.8%), Tandjilé (88.9%), Wadi Fira (88.9%), Mandoul (86.2%), Ennedi (85.3%), Mayo Kebbi Est (84.2%), Logone Occidental (80.4%), all in Chad require critical prioritization.

**Figure 2:**
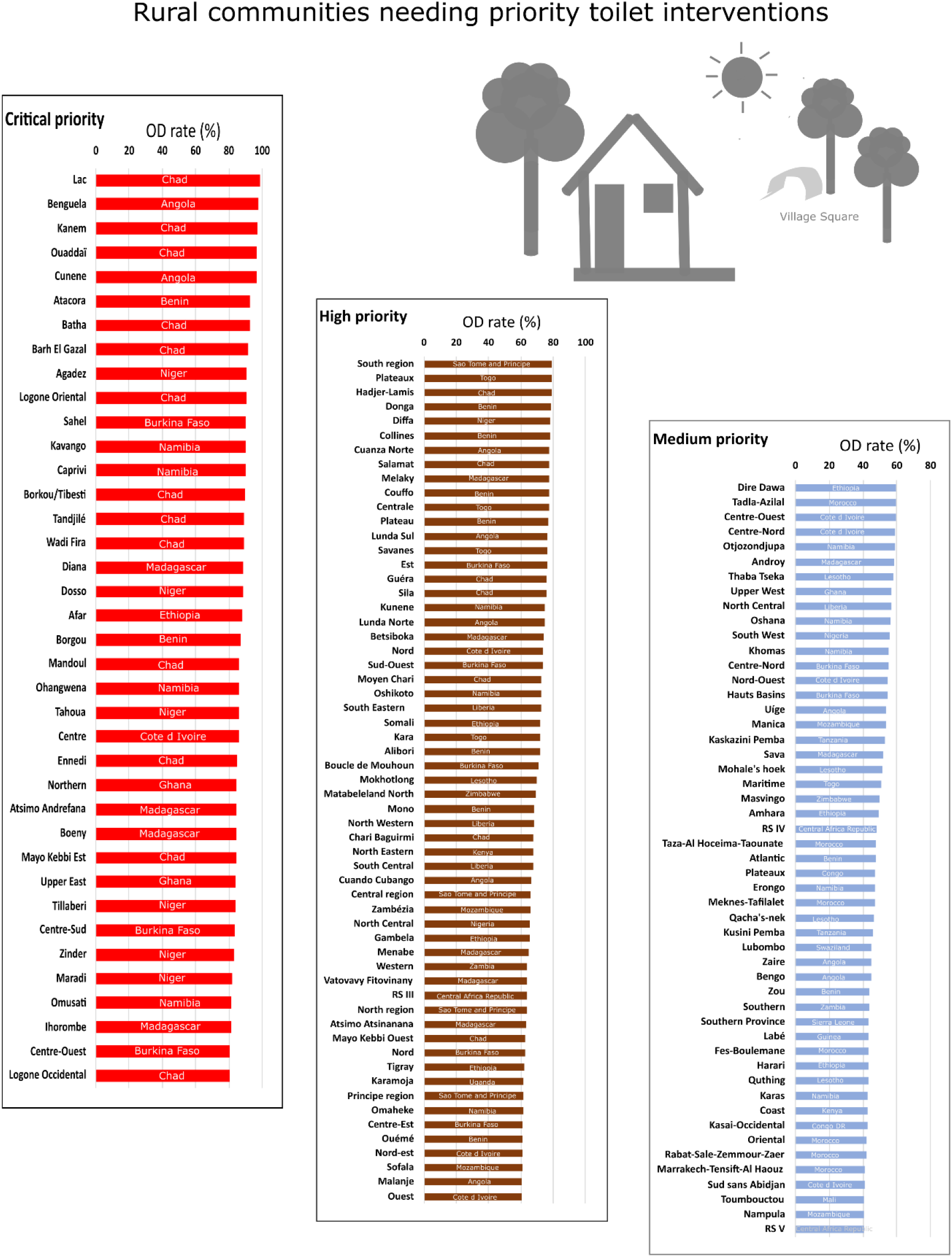
Rural communities in Africa needing priority interventions against open defecation.

Other regions that need critical rural prioritising are **Angola** (*Benguela, Cunene, Bamibe, Huila, Cuanza Sul*), **Namibia** (*Kavango, Caprivi, Ohangwena, Omusati*), **Madagascar** (*Diana, Atsimo Andrefana, Boeny, Ihorombe*), **Benin** (*Atacora, Borgou*), **Burkina Faso** (*Sahel, Centre-Sud, Centre-Quest*), **Niger** (*Agadez, Dosso, Tahoua, Tillaberi, Zinder, Maradi)*, **Cote d’Ivoire** (Centre), **Ethiopia** (*Afar*), and **Ghana** (Northern, Upper East). Similarly, 59 out of 412 sub-national regions in this study have a high OD rate in rural areas (Figure 2).

Countries with one or more regions with critical open defecation rates by residence are Angola, Benin, Burkina Faso, Chad, Cote d’Ivoire, Ethiopia, Ghana, Madagascar, Namibia, Niger, and Sao Tome and Principe (Table 5). Irrespective of residence, all the Sao Tome and Principe regions have sanitation challenges requiring at least one of the open defecation priority systems. The datasets analysed were from the DHS Program (DHS, 2022).

**Table 5:** Summary of countries that have regions with critical open defecation rates.

| Countries with geographical regions in critical state by number of regions |  |  |  |  |  |  |  | Priority<br>Critical (≥80%)<br>High (60-≤80%)<br>Medium (40-≤60%) |
| --- | --- | --- | --- | --- | --- | --- | --- | --- |
| Countries | Total region | Urban |  |  | Rural |  |  |  |
|  |  | Critical | High | Medium | Critical | High | Medium |  |
| Angola | 18 | 0 | 0 | 0 | 5 | 5 | 3 |  |
| Benin | 12 | 1 | 0 | 7 | 2 | 7 | 2 |  |
| Burkina Faso | 13 | 0 | 1 | 0 | 3 | 5 | 2 |  |
| Chad | 21 | 0 | 2 | 6 | 13 | 7 | 0 |  |
| Cote d Ivoire | 11 | 0 | 0 | 0 | 1 | 3 | 4 |  |
| Ethiopia | 11 | 0 | 0 | 0 | 1 | 3 | 3 |  |
| Ghana | 10 | 0 | 0 | 1 | 2 | 0 | 1 |  |
| Madagascar | 22 | 0 | 0 | 3 | 4 | 5 | 2 |  |
| Namibia | 13 | 0 | 1 | 4 | 4 | 3 | 5 |  |
| Niger | 8 | 0 | 0 | 0 | 6 | 1 | 0 |  |
| Sao T&P | 4 | 1 | 0 | 3 | 0 | 4 | 0 |  |

Table 6 below shows that the availability of toilets is directly proportional to the wealth index. Within countries with critical sanitation challenges, the poor (i.e. the poorer and the poorest) account for about 60.8% of the total open defecation compared to 14.7% among the rich (i.e. the richer and the richest). Similarly, the higher the level of education, the lower the possibility of practicing open defecation. The datasets were from the DHS Program (DHS, 2022). A flush toilet is common in urban areas while open defecation is common in rural areas. However, the latrine is used in both urban and rural areas.

**Table 6:** Descriptives and regression case processing summary.

| **Descriptives statistics for regressions** |  |  |  |  |  |  |
| --- | --- | --- | --- | --- | --- | --- |
| **Wealth index** |  |  |  |  |  |  |
| **Toilet type** | Poorest | Poorer | Middle | Richer | Richest | Total |
| Flush | 1166 | 5787 | 12462 | 21156 | 43240 | 83811 |
| Latrine | 16751 | 36148 | 47838 | 55788 | 73088 | 229613 |
| No toilet | 120742 | 80829 | 60043 | 38245 | 6879 | 306738 |
| Total | 138659 | 122764 | 120343 | 115189 | 123207 | 620162 |
| **Highest educational level** |  |  |  |  |  |  |
| **Toilet type** | No education | Primary | Secondary | Higher | Don't know | Total |
| Flush | 23493 | 24775 | 26001 | 6537 | 360 | 81166 |
| Latrine | 86097 | 61328 | 31991 | 5461 | 361 | 185238 |
| No toilet | 172526 | 72130 | 19329 | 807 | 274 | 265066 |
| Total | 282116 | 158233 | 77321 | 12805 | 995 | 531470 |
| **Type of place of residence** |  |  |  |  |  |  |
| **Toilet type** | Urban | Rural | Not applicable |  |  | Total |
| Flush | 69850 | 13961 |  |  |  | 83811 |
| Latrine | 102853 | 126760 |  |  |  | 229613 |
| No toilet | 39625 | 267113 |  |  |  | 306738 |
| Total | 212328 | 407834 |  |  |  | 620162 |

Logistics regression analyses of the eleven countries with critical priority regions are presented in Figure 5. Figure 5a below indicates that the poorest in these countries are more likely to be without a toilet and practice open defecation. After adjusting for covariates, the poorest are 43 times more likely to practice open defecation than the richest (95% CI *= 42.443 – 45.290*). Even the wealthier people are about five times more likely to practice open defecation than the richest (95% CI = *5.039 – 5.355*).

Open defecation rate in seven most populous countries in Africa namely Nigeria, Ethiopia, Egypt, Democratic Republic of Congo (DRC), South Africa, Tanzania and Kenya are presented in figures 3 and 4 using datasets analysed were from the DHS Program and World Bank population ranking (DHS, 2022; World Bank, 2023).

**Figure 3:**
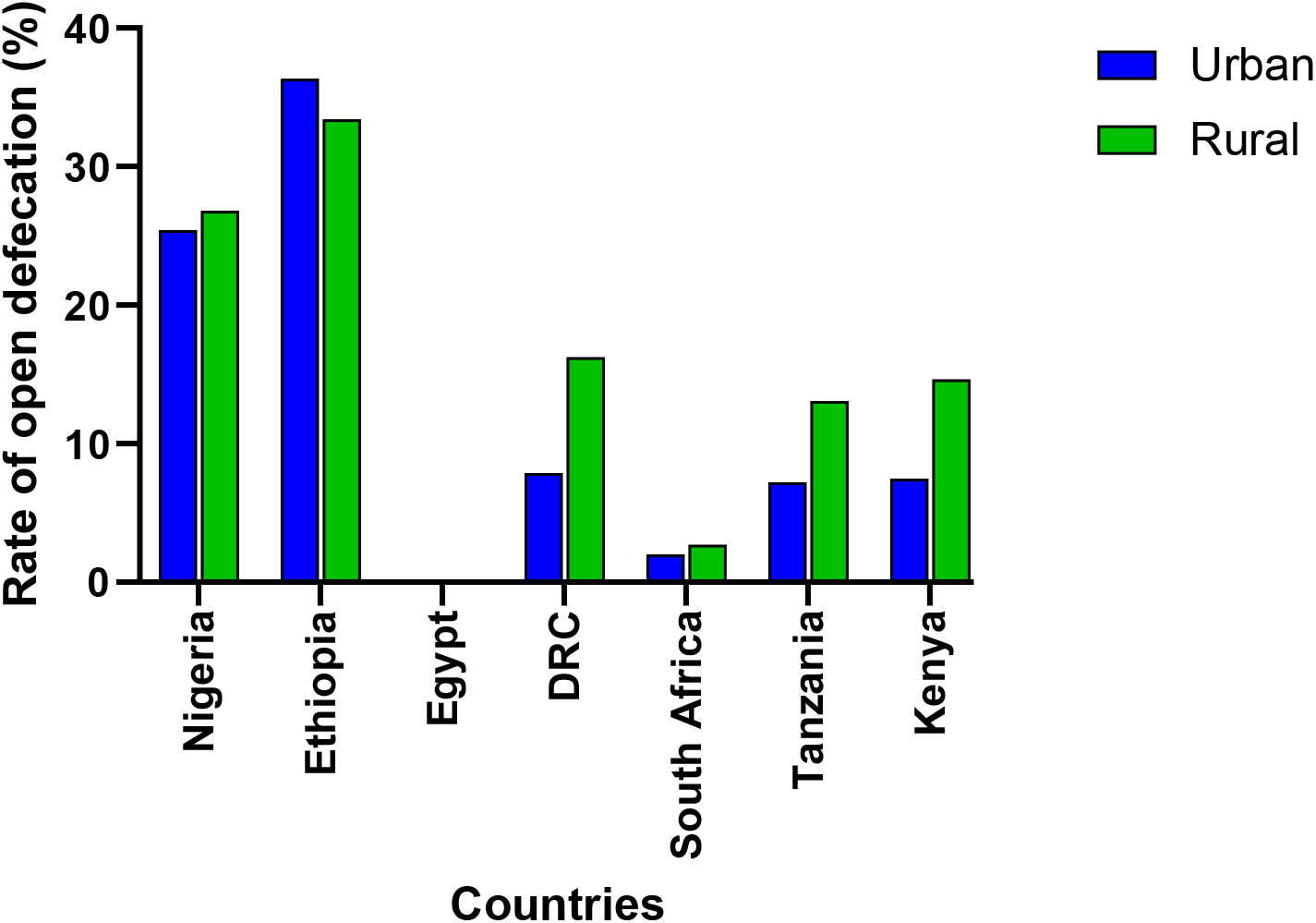
Open defecation rate in seven most populous countries in Africa.

**Figure 4:**
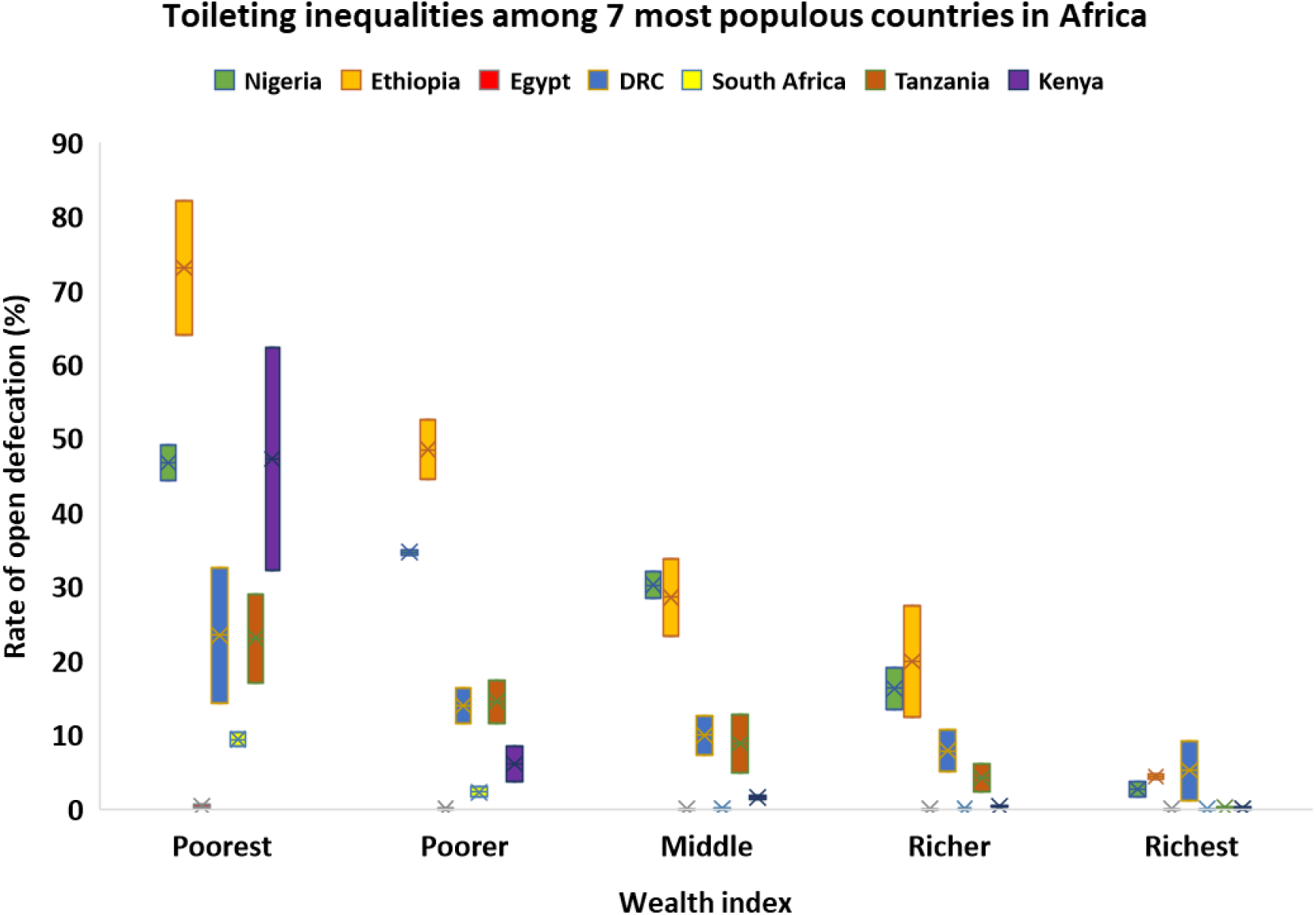
Toileting inequalities among Africa’ *big seven*.

**Figure 5:**
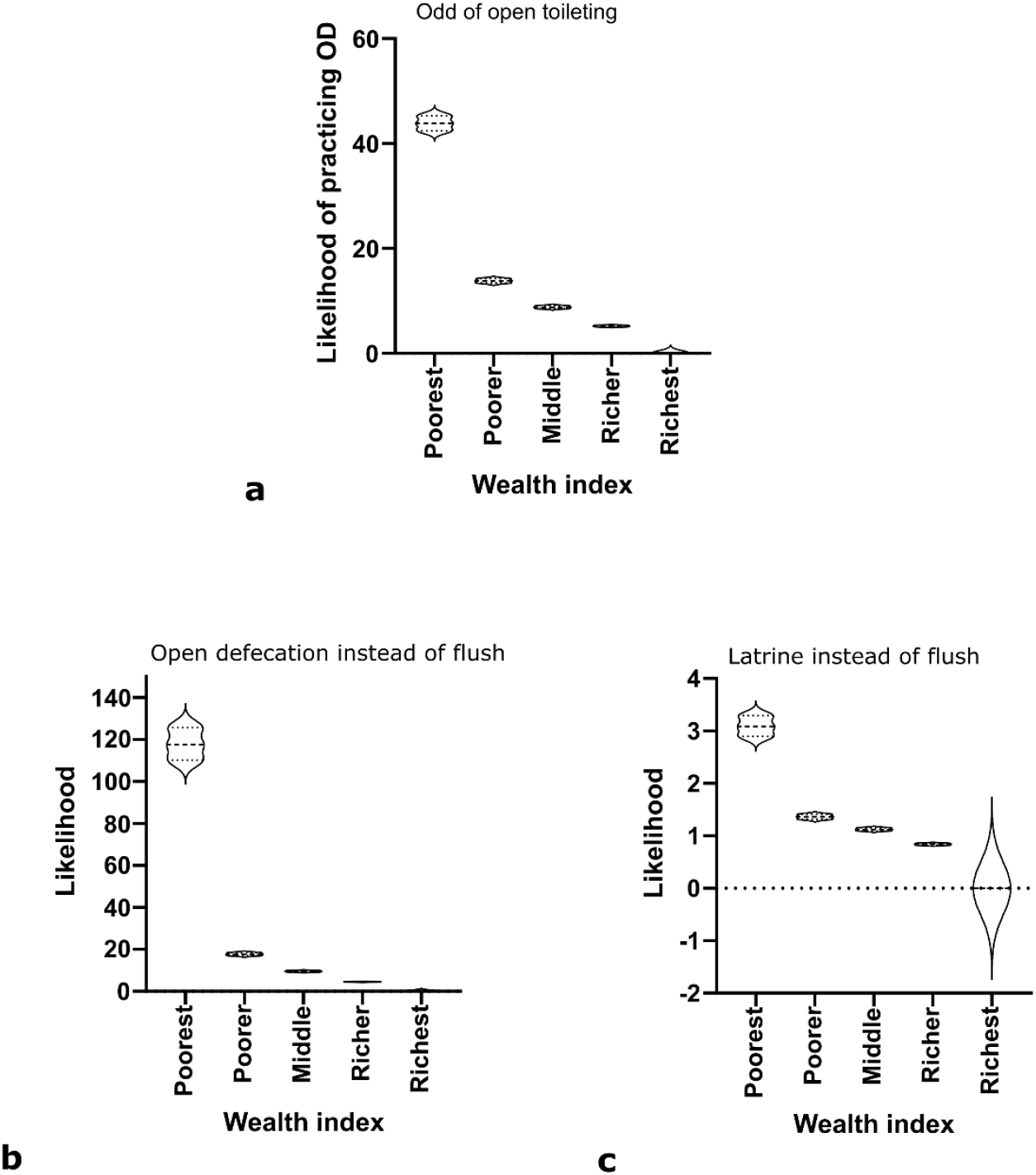
Logistic regression of African regions in critical state. (a) Likelihood of practicing open defecation (b) Likelihood of defecating in the open instead of flush toilet (c) Likelihood of using latrine instead of flush toilet.

The length of the boxes in figure 4 indicates the margin in rural vs urban areas. Among these seven populous Africa countries, open defecation rate is lowest in Egypt. Egypt peak OD practice is among the most impoverished rural residents, and the rate is about 0.6%. On the other hand, OD is highest in Ethiopia, especially among the poorest residents in rural (82%). Generally, the poorest in urban areas have a reduced open defecation rate than the poorest in rural areas. The margin among the poorest is vast in Kenya, Ethiopia, and Democratic Republic of Congo (DRC). Among the richest in DRC, the place of residence widens the open defecation rate.

For African regions with critical state, the poorest are 43 times more likely to defecate in the open than the richest (Figure 5a). However, beyond the binary of had toilet and no toilet, multinomial categorisation (flush, latrine, and no toilet) revealed how vast toileting inequalities could be (Figures 5b and 5c). For instance, figure 5c indicates that among people with no toilet, the poorest are 117 times more likely to practice open defecation (95% CI = *110.172– 125.786*) than the richest. The regression analyses indicate that in areas with critical OD practice, wealth index is a vital factor that promotes this sanitation indicator.

## DISCUSSION

### Leadership on Open Defecation

In 2008, the African Union (AU) pledged to improve water and sanitation status during her 11th ordinary session in Sharm el-Sheikh, Egypt. This pledge acknowledged the crucial role of water and sanitation in promoting social, economic, and environmental development for member countries and the entire African continent (AU, 2008). Albeit, five years after, these pledges were not reflected in the continent’s big agenda. In January 2013, the AU summit adopted Agenda 2063 – The Africa We Want. The 50-year-long agenda is intended to be Africa’s blueprint and master plan for sustainable development and the continent’s economic growth (AU, 2021; NEPAD, 2020). Commitment towards ending open defecation or safe and hygienic toilets for all is not made in the “The voices of the African people” 63 statements (AU, 2015). At first while AU attempted to link its agenda with the global sustainable plan, none of the goals and priority areas clearly addressed sanitation (AU, 2021). The first implementation report by the pan-Africa body does not contain sanitation (NEPAD, 2020). However, the 2022 implementation report had sanitation as an indicator in one of the priority areas of goal one (NEPAD, 2022). Sadly, sanitation is an afterthought within the continent’s own agenda. The present analysis (Table 2) and low progress and commitment to sanitation suggest that open defecation may continue in Africa until 2030 or beyond.

Notwithstanding, recent times have seen a renewed continental effort on sanitation. Firstly, in 2015, Ngor Declaration was made by African Ministers Council on Water (AMCOW), a specialised committee for water and sanitation in AU. The declaration calls for universal access to adequate and sustainable sanitation and hygiene services and for eliminating open defecation by 2030 (AMCOW, 2019). AMCOW seeks to respond to the need to reform sanitation policies in the continent to reduce the number of people without access to sanitation services and improve access to safely managed sanitation. A recent output of the declaration is the African Sanitation Policy Guidelines (ASPG), designed to guide African governments in reviewing, revising, and developing sanitation policy and its implementation strategy (AMCOW, 2021). While this is a laudable effort, implementing the country-specific sanitation policy may present a bottleneck.

Poverty or wealth index is a factor linked to the incessant practice of OD (Figures 4 and 5). Nigeria’s position is crucial in determining the success of open defecation-free (ODF) Africa because of its current poverty rate of 43%, which is about 89.8 million people (WPC, 2021). Africa will require a concerted effort to be ODF since 263 million people across Nigeria, Ethiopia, Niger, DR Congo, Burkina Faso, Chad, Angola, Madagascar, Kenya, and Cote d’Ivoire live in extreme poverty (Table 2, WPC, 2021). These ten countries with toilet emergency account for about 54% of Africans in extreme poverty, and the current trend suggests that ODF status in these countries may not be achieved by 2030. Albeit this may be readily implemented through the combined efforts of the government and a broad range of stakeholders in building a solid foundation for access to sanitation (UNICEF and WHO, 2020).

### Wealth and Health

The work highlights that the poverty line is still a determining factor why people defecate in the open, especially in areas where the OD rate is ≥80% (Figure 5). After adjusting for education and place of residence covariates, the poorest are 43 times more likely to practice open defecation than the richest (95% CI = *42.443 – 45.290*). Nigeria and Ethiopia are core drivers of the African population practicing open defecation (Figures 3 and 4). Some years back toileting intervention in India failed partly because the poorest were provided with toilets without inspiring behavioural change about sanitation (Singh and WSP, 2007). There are similar reports of the negative influence of lower socioeconomic status on open defecation practice in Kenya (Busienei *et al.,* 2019), India (Vyas *et al.,* 2019), Nepal (Bhatt *et al.,* 2019), and Ghana (Adzawla *et al.,* 2020). Truly, behavioural change is essential as toilet provision does not always equate to the end of open defecation. Notwithstanding, positive wealth status can stimulate positive behavioural change about sanitation. Studies have shown that income is associated with better health, and wealth affects health (Braveman *et al.,* 2010; NCHS, 2012; Pollack *et al.,* 2013; Gordon *et al.,* 2020). Perhaps, more Africentric or country-specific studies need to be carried out on socioeconomic status and health.

### Toileting issues in West Africa and the high burden of antimicrobial resistance

Historically, open defecation has been endemic to West Africa, and the trend subsists in the region (Tables 1 – 4). OD practice is prevalent in West African countries. For instance, Nigeria has the highest number of people in the continent practising OD (Table 2). Other Countries that share boundaries with her, viz Niger, Chad, and Benin, also struggle to end open defecation (Table 4). Urban open defecation is at a critical-priority level in the *Atacora* region of Benin and the *Est* region of Burkina Faso (Figure 1).

Furthermore, out of the 42 regions with a critical rural OD rate, 27 (64%) are West African sub-national (Figure 2). *Atacora*, a largely rural area, is the poorest in Benin in non-monetary poverty (World Bank, 2009). *Atacora* lies in northwestern Benin and shares a border with the *Est* region of Burkina Faso. Both regions share a forested, rugged mountainous range and a sanitation dilemma. Sanitation bottlenecks in these regions include harmful social norms and the unwillingness of households to build, use and manage the toilets. Because of the mountainous terrain, the regions also face hydrogeological challenges that impede borehole drilling and the use of flush toilets (UNICEF Burkina Faso, 2019). CLTS is being implemented by local enterprises that construct toilets, and villages get ODF status and certification. Sustainability checks in Côte d’Ivoire, Benin and Ghana reveal that there is a high rate of post-ODF slippage, and this is a result of a decline in the use of latrine toilets. The decrease in the use of toilets was reported to be due to the use of low-quality hardware for toilet construction, low know-how capabilities of the local monitoring committee, limited involvement of local authorities in programme implementation, and uncompetitive commercial sanitation market (Jiménez *et al.,* 2017).

West African regional bloc, the Economic Community of West African States (ECOWAS), formed its health arm in 1987 to protect member states’ health. However, since 2000, when West Africa Health Organisation (WAHO) became operational, there has been no strong leadership in sanitation until recently with ECOWAS Vision 2050 (WAHO, 2020; ECOWAS, 2022). It is not enough to have a few mentions of words like ’sanitation’ or ’WASH’ in policies, rather, statements need to be backed with commitments, initiatives and influence on member states toward ODF. For instance, the press release by WAHO to mark 2020 *World Toilet Day* does not highlight the degree of the problems of open defecation in West Africa. By referring to only global statistics instead of succinct regional facts and implications, it appears WAHO does not grasp the magnitude of OD in its region. The open defecation challenge in ECOWAS is beyond the annual November 19^th^ statement, press release and one-off ceremony. It is noteworthy that less than a decade to 2030, ECOWAS and its specialised health agency WAHO do not seem to have a solid road map on ODF or any WASH component.

The World Health Organisation has recognised antimicrobial resistance as a priority’ global health epidemic’ and ’development threat’ facing humanity (WHO, 2021). Of all the regions and sub-regions of the world, west Africa has the highest all-age death caused by drug resistance (Murray *et al.,* 2022). Furthermore, it is confirmed that antibiotics residues are present in faecal wastes, which increases the burden of AMR on release to the environment, a predominant case in West Africa (Hendriksen *et al.,* 2019; Wilkinson *et al.,* 2022). Indeed, west Africa has a huge and significant toilet problem that could be driving the high burden of AMR in the region and worldwide (Murray *et al.,* 2022).

### Water sanitation and hygiene financing and the lesson from Egypt and South Africa

As shown in Table 3, Egypt has been leading Africa in successful access to sanitation, which WaterAid also reported (2015) and recent AU agenda 2063 (NEPAD, 2022). Unlike the OD rate of 0.7% presented in Table 3, WaterAid reported that Comoros has a challenge with access to sanitation (64% of the population). It is to be noted that the result used the year 2012 datasets, which are the latest publicly available datasets for Comoros. WaterAid may have used more recent data that is unavailable as at the time of data analyses in 2021. Moreso, it reported a percentage (%) of the population without access to sanitation, not specifically *% of OD*. A similar disparity is observed for Rwanda, Cameroon, and a few other countries with old data. Despite the successes of Egypt, with a low rate of open defecation, because most households have latrines or flush toilets, wastewater management is still a challenge (Harmsen *et al.,* 2014). This challenge is common in rural Upper Egypt or Nile Delta because of the high population density (Harmsen *et al.,* 2014; World Bank, 2015). The country has used a CLTS-type approach for sanitation and waste management (IIED, 2010). More importantly, the secret to eliminating open defecation is an investment in WASH. Between July 2014 till July 2020, Egypt invested EGP 174 billion to ensure equitable and sustainable water and sanitation (about USD 11 billion) (Mohammed, 2020). Likewise at the end of apartheid in South Africa, the government committed to financing the implementation of WASH policies (GoSA and UNDP, 2005). Consequently, open defecation dropped from 12.4% in 2000 to 1.4% in 2017 (WHO/UNICEF JMP, 2021). Table 3 presents 2.6% for South Africa, a slightly higher rate. Ensuring proper sanitation goes beyond the construction of toilets, as these structures will inevitably reach their capacity and potentially become inoperable. To overcome this and offer a long-term maintenance plan, there was an early commitment to WASH research and community engagement in the middle-income country of South Africa (Still *et al.,* 2012; Ntaro *et al.,* 2022). Government-led funds in Africa will play a crucial role in promoting scientific knowledge and practical solutions for the efficient management of faecal wastes and eradicating open defecation.

### Conclusions

Open defecation is a significant challenge that pose health and socio-economic challenges in developing countries. To address this and other developmental challenges, the UN created the MDGs, followed by the global SDGs targets to enhance the progress made during MDGs. However, less than seven years to the end of SDGs 2030, Africa is still behind on many indices, particularly in using safely managed toilets. In this context, this study investigates the practice, status, and sociodemographic drivers of open defecation in African countries. The study used publicly available DHS Program datasets of over eight million respondents across 40 countries to achieve these objectives.

The findings reveal that most African countries historically and presently practice open defecation, out of which ten countries are projected by year 2030 to have around 247 million people without safely managed toilets. These findings indicate the urgent need for improved leadership and efforts from the African Union, sub-regional organizations, and countries in implementing, monitoring, and evaluating sanitation policies and programmes. Likewise, the findings highlight that poverty is a significant determining factor of open defecation practice. Poor people are significantly more likely to practice open defecation than the rich. Furthermore, the study reveals disparities in access to toilets in rural areas, exacerbating the problem of open defecation. Therefore, empowering rural communities and providing community-led water, sanitation, and hygiene services are crucial to achieving open defecation-free Africa. It is essential to highlight that a nexus exist among the SDGs with a strong link between SDGs 6, 1, 3, 4 and 11. Clean water and sanitation supports the promotion of good health and wellbeing and ensure sustainable cities and communities, which reinforces the need for quality education that leads to the reduction of poverty.

Policymakers must prioritise interventions to end open defecation in Africa, focusing on regions and communities where the problem is most severe and requires significant resources. Such interventions should be informed by data, and trans-national organizations and country governments must work together to improve leadership and efforts in implementing, monitoring, and evaluating sanitation policies and interventions. Likewise, because of the high burden of AMR in Africa and the growing reports of drug-resistant pathogens in faeces, steps must be taken to address the threat of AMR by ending open defecation and monitoring interventions and burdens through the lens of One Health. This should include holistic multisectoral approaches to increasing surveillance and stewardship intermediations in those priority communities, regions, and countries identified in this study.

The limitation of the study using DHS program datasets includes not accounting for some areas or populations that may be underrepresented due to a multistage sampling approach. In addition, the survey relies on self-reported data, which can be subject to biases such as social desirability or recall bias. Likewise, the data may not reflect the most recent changes or trends since DHS surveys are typically conducted every 5-10 years. Finally, data unavailability results in the unintended exclusion of certain African countries. Despite these limitations, DHS data remains a valuable source for policymakers, development partners and researchers working to improve the wellbeing of developing countries. Future research should address these limitations and provide a more comprehensive health survey that captures variables on global health concerns such as AMR. Nonetheless, this study provides valuable insights into the persistent problem of open defecation in Africa and highlights the urgent need for effective interventions to address this issue. In summary, interventions to end open defecation in Africa should be multidisciplinary and informed by data, concentrating on communities and regions where the problem is most severe and requires significant resources.

### Recommendations

Based on the findings of this research, some policy recommendations for addressing the issue of open defecation in Africa should include first, targeted and data-driven interventions to facilitate needs-based approach that focus on regions and communities in urgent need. The findings of the study highlight the critical, high, and medium priority areas, which can guide the allocation of resources and intervention strategies.

Secondly, there should be emphasis on community-led total sanitation. The CLTS has been relatively successful in reducing open defecation in many parts of the world. The use of appropriate policy has the potential to lead to a high uptake of the model in several African countries to help combat open defecation in their communities.

Thirdly, it is important to address poverty and inequality to improve sanitation. Inequalities in toileting were observed among individuals living in poverty or in rural areas. Lack or inadequate sanitation infrastructure is a significant contributor to open defecation in Africa. Governments should invest in the improvement of sanitation infrastructure, including the construction of public toilets and sanitation facilities in rural areas backed with community support and leadership. Governments and regional bodies through dedicated agencies should create a special fund or allocate a percentage of the government budget toward sanitation infrastructure. Interventions should target the root causes of poverty and inequality, which drive open defecation.

Fourthly, it is essential to understand and promote positive behavioural change on sanitation. Insights from behavioural studies can be used in diagnosis and development interventions that are essential for combating open defecation. Governments across Africa should promote behavioural change through awareness campaigns, education, and social marketing to encourage people to use toilets and improve hygiene practices.

Fifthly, important surveys such as demographic and health survey which is widely deployed in developing countries should incorporate variables that will assist in surveillance of antimicrobial resistance, including the collection and analyses of environmental samples or faecal samples from selected sites and household during the survey.

Finally, collaborations and partnerships with governments, non-governmental organisations, funding bodies and other stakeholders is crucial to share knowledge, resources, and expertise to end open defecation in Africa. The involvement of the private sector and alternative funding mechanism is vitally important in promoting sanitation infrastructure development and investment in sanitation programs.

## Supporting information

Regressions

Datasets characteristics

## Data Availability

The data used in this study are publicly available and accessed at https://dhsprogram.com/, explored by country, survey type and year.

