## Supplementary material for "Analyses of health surveys indicates regions of priority to eliminate open defecation in Africa and implication for antimicrobial resistance burden": Regressions

**SUPPLEMENTARY DATA**

**Supplementary 2**

|  |  | Model 1 (unadjusted) | | | | Model 2 | | | | Model 3 | | | |
| --- | --- | --- | --- | --- | --- | --- | --- | --- | --- | --- | --- | --- | --- |
| Toilet ^a^ | Covariates/Factor | Sig. | AOR | 95% Confidence Interval | | Sig. | AOR | 95% Confidence Interval | | Sig. | AOR | 95% Confidence Interval | |
|  |  |  |  | Lower | Upper |  |  | Lower | Upper |  |  | Lower | Upper |
| No toilet/Open defecation | Education |  |  |  |  | 0.000 | 0.637 | 0.632 | 0.643 | 0.000 | 0.694 | 0.687 | 0.700 |
|  | Residence |  |  |  |  |  |  |  |  | 0.000 | 3.157 | 3.107 | 3.207 |
|  | Poorest | 0.000 | 110.597 | 107.459 | 113.826 | 0.000 | 89.538 | 86.790 | 92.373 | 0.000 | **43.844** | **42.443** | **45.290** |
|  | Poorer | 0.000 | 31.972 | 31.121 | 32.846 | 0.000 | 27.680 | 26.881 | 28.502 | 0.000 | 13.830 | 13.411 | 14.262 |
|  | Middle | 0.000 | 16.583 | 16.145 | 17.033 | 0.000 | 15.032 | 14.601 | 15.475 | 0.000 | 8.791 | 8.530 | 9.059 |
|  | Richer | 0.000 | 8.313 | 8.090 | 8.542 | 0.000 | 8.107 | 7.872 | 8.349 | 0.000 | 5.195 | 5.039 | 5.355 |
| Model Fitting Criteria | AIC | 228670.247 | | | | 210706.191 | | | | 234671.735 | | | |
|  | BIC | 228681.596 | | | | 210717.385 | | | | 234682.929 | | | |
|  | -2log likelihood | 228668.247 | | | | 210704.191 | | | | 234669.735 | | | |

Supplementary 2: Binomial logistic regression of African countries that have regions in a critical state.

^a^ The reference category is: Had toilet.

**Supplementary 3**

|  |  | Model 1 (unadjusted) | | | | Model 2 | | | | Model 3 | | | |
| --- | --- | --- | --- | --- | --- | --- | --- | --- | --- | --- | --- | --- | --- |
| Toilet ^a^ | Covariates/Factor | Sig. | AOR | 95% Confidence Interval | | Sig. | AOR | 95% Confidence Interval | | Sig. | AOR | 95% Confidence Interval | |
|  |  |  |  | Lower | Upper |  |  | Lower | Upper |  |  | Lower | Upper |
| Latrine Toilet | Education |  |  |  |  | 0.000 | 0.690 | 0.684 | 0.696 | 0.000 | 0.739 | 0.733 | 0.746 |
|  | Residence |  |  |  |  |  |  |  |  | 0.000 | 4.240 | 4.142 | 4.339 |
|  | Poorest | 0.000 | 8.499 | 8.000 | 9.030 | 0.000 | 7.150 | 6.719 | 7.609 | 0.000 | 3.088 | 2.898 | 3.290 |
|  | Poorer | 0.000 | 3.695 | 3.586 | 3.809 | 0.000 | 2.972 | 2.880 | 3.067 | 0.000 | 1.364 | 1.318 | 1.411 |
|  | Middle | 0.000 | 2.271 | 2.219 | 2.324 | 0.000 | 1.847 | 1.803 | 1.893 | 0.000 | 1.123 | 1.094 | 1.153 |
|  | Richer | 0.000 | 1.560 | 1.530 | 1.591 | 0.000 | 1.229 | 1.203 | 1.255 | 0.000 | 0.842 | 0.823 | 0.861 |
| No toilet/Open defecation | Education |  |  |  |  | 0.000 | 0.480 | 0.475 | 0.485 | 0.000 | 0.546 | 0.540 | 0.553 |
|  | Residence |  |  |  |  |  |  |  |  | 0.000 | 9.719 | 9.484 | 9.960 |
|  | Poorest | 0.000 | 650.909 | 611.144 | 693.261 | 0.000 | 451.384 | 422.953 | 481.726 | 0.000 | 117.720 | 110.172 | 125.786 |
|  | Poorer | 0.000 | 87.796 | 84.619 | 91.092 | 0.000 | 63.461 | 61.037 | 65.982 | 0.000 | 17.746 | 17.028 | 18.495 |
|  | Middle | 0.000 | 30.286 | 29.334 | 31.268 | 0.000 | 23.386 | 22.601 | 24.198 | 0.000 | 9.569 | 9.231 | 9.918 |
|  | Richer | 0.000 | 11.363 | 11.022 | 11.715 | 0.000 | 9.362 | 9.060 | 9.673 | 0.000 | 4.537 | 4.383 | 4.696 |
| Model Fitting Criteria | AIC | 245290.903 | | | | 234697.322 | | | | 276408.369 | | | |
|  | BIC | 245313.578 | | | | 234719.689 | | | | 276430.736 | | | |
|  | -2log likelihood | 245286.903 | | | | 234693.322 | | | | 276404.369 | | | |

Supplementary 3: Multinomial logistic regression of African countries that have regions in a critical state.

^a^ The reference category is: Flush toilet.
